# Adiposity, metabolites, and colorectal cancer risk: Mendelian randomization study

**DOI:** 10.1101/2020.03.19.20031138

**Authors:** Caroline J. Bull, Joshua A. Bell, Neil Murphy, Eleanor Sanderson, George Davey Smith, Nicholas J. Timpson, Barbara L. Banbury, Demetrius Albanes, Sonja I. Berndt, Stéphane Bézieau, D Timothy T. Bishop, Hermann Brenner, Daniel D. Buchanan, Andrea Burnett-Hartman, Graham Casey, Sergi Castellví-Bel, Andrew T. Chan, Jenny Chang-Claude, Amanda J. Cross, Albert de la Chapelle, Jane C. Figueiredo, Steven J. Gallinger, Sue M. Gapstur, Graham G. Giles, Stephen B. Gruber, Andrea Gsur, Jochen Hampe, Heather Hampel, Tabitha A. Harrison, Michael Hoffmeister, Li Hsu, Wen-Yi Huang, Jeroen R. Huyghe, Mark A. Jenkins, Corinne E. Joshu, Temitope O. Keku, Tilman Kühn, Sun-Seog Kweon, Loic Le Marchand, Christopher I. Li, Li Li, Annika Lindblom, Vicente Martín, Anne M. May, Roger L. Milne, Victor Moreno, Polly A. Newcomb, Kenneth Offit, Shuji Ogino, Amanda I. Phipps, Elizabeth A. Platz, John D. Potter, Conghui Qu, J. Ramón Quirós, Gad Rennert, Elio Riboli, Lori C. Sakoda, Clemens Schafmayer, Robert E. Schoen, Martha L. Slattery, Catherine M. Tangen, Kostas K. Tsilidis, Cornelia M. Ulrich, Franzel JB. van Duijnhoven, Bethany Van Guelpen, Kala Visvanathan, Pavel Vodicka, Ludmila Vodickova, Hansong Wang, Emily White, Alicja Wolk, Michael O. Woods, Anna H. Wu, Peter T. Campbell, Wei Zheng, Ulrike Peters, Emma E. Vincent, Marc J. Gunter

## Abstract

**Importance:** Evidence on adiposity altering colorectal cancer (CRC) risk differently among men and women, and on metabolic alterations mediating effects of adiposity on CRC, is unclear.

**Objective:** To examine sex- and site-specific associations of adiposity with CRC risk, and whether adiposity-associated metabolites explain associations of adiposity with CRC.

**Design:** Two-sample Mendelian randomization (MR) study.

**Setting:** Genetic variants from expanded genome-wide association studies of body mass index (BMI) and waist-to-hip ratio (WHR, unadjusted for BMI; N=806,810), and 123 metabolites (mostly lipoprotein subclass-specific lipids) from targeted nuclear magnetic resonance metabolomics (N=24,925), were used as instruments. Sex-combined and sex-specific MR was conducted for BMI and WHR with CRC risk; sex-combined MR was conducted for BMI and WHR with metabolites, for metabolites with CRC, and for BMI and WHR with CRC adjusted for metabolite classes.

**Participants:** 58,221 cases and 67,694 controls (Genetics and Epidemiology of Colorectal Cancer Consortium; Colorectal Cancer Transdisciplinary Study; Colon Cancer Family Registry).

**Main outcome measures:** Incident CRC (overall and site-specific).

**Results:** Among men, higher BMI (per 4.2 kg/m^2^) was associated with 1.23 (95%-confidence interval (CI)=1.08, 1.38) times higher CRC odds (inverse-variance-weighted (IVW) model); among women, higher BMI (per 5.2 kg/m^2^) was associated with 1.09 (95%-CI=0.97, 1.22) times higher CRC odds. Higher WHR was more strongly associated with CRC risk among women (IVW-OR=1.25, 95%-CI=1.08, 1.43 per 0.07-ratio) than men (IVW-OR=1.05, 95%-CI=0.81, 1.36 per 0.07-ratio). BMI or WHR was associated with 104 metabolites (false-discovery-rate-corrected P≤0.05) including low-density lipoprotein (LDL) cholesterol, but these metabolites were generally unassociated with CRC in directions consistent with mediation of adiposity-CRC relations. In multivariable MR, associations of BMI and WHR with CRC were not attenuated following adjustment for representative metabolite classes – e.g. the univariable IVW-OR of BMI for CRC was 1.12 (95%-CI=1.00, 1.26), and 1.11 (95%-CI=0.99, 1.26) adjusting for LDL lipids.

**Conclusions and relevance:** Our results suggest that higher BMI more greatly raises CRC risk among men, whereas higher WHR more greatly raises CRC risk among women.

Adiposity was associated with numerous metabolic alterations, but none of these alterations explained associations between adiposity and CRC. More detailed metabolomic measures are likely needed to clarify mechanistic pathways.

## Introduction

Colorectal cancer (CRC) is one of the most commonly diagnosed cancers among adults globally (1-3). Obesity is viewed as a likely cause of CRC by the International Agency for Research on Cancer (IARC), the American Institute for Cancer Research (AICR), and the World Cancer Research Fund (WCRF) (3, 4), based largely on positive associations between adiposity and CRC risk from observational epidemiology. Mendelian randomization (MR) studies, which use genetic variants as instruments (proxies) for adiposity given their randomly allocated and fixed nature (5), further support causality (6-8). Despite this growing consensus, it remains unclear whether the effect of adiposity on CRC risk differs among men and women, whether the relationship varies by CRC subsite, and what the underlying biological mechanisms are. These are important to clarify given the on-going obesity epidemic and difficulties in reducing adiposity itself (9, 10).

Observationally, body mass index (BMI) relates more strongly to CRC risk among men and waist-to-hip ratio (WHR) relates similarly to CRC risk among men and women (11). However, recent MR studies suggest that higher BMI more greatly raises CRC risk among women, while higher WHR more greatly raises CRC risk among men (6, 7). Whether these MR estimates are robust is unclear because they were based on relatively small sample sizes, genetic instruments that were not sex-specific, and genetic instruments for WHR that were conditioned on BMI – all potential sources of bias (12-16). Adiposity alters systemic metabolism (17-19), but evidence for effects of adiposity-altered metabolites on CRC is scarce. One MR study suggested that total cholesterol raises CRC risk (20), while others suggested no effect of blood glucose (21) and mixed support for fatty acids (22). Overall, the scope of metabolic traits examined has also been narrow. Targeted metabolomics allows deeper phenotyping at large scale (23), and its recent integration with genotype data (24) enables us to examine associations of metabolites with CRC using MR. Expanded genotype data for CRC is also available (25), affording a sample size six-times larger than used in previous MR studies (58,221 cases, 67,694 controls).

This study has two aims. First, we aimed to better estimate sex-specific effects of adiposity on CRC risk using two-sample MR. We examined associations of BMI and WHR with CRC risk using expanded GWAS data and genetic instruments for exposures that were sex-specific and were not mutually conditioned, to reduce bias (12-16). Second, we aimed to identify potential metabolic mediators of effects of adiposity on CRC risk using two-step MR (by examining associations of BMI and WHR with metabolites, and of BMI-or WHR-related metabolites with CRC risk) and multivariable MR (by adjusting associations of BMI and WHR with CRC for representative metabolites).

## Methods

### Study design

We used two-sample MR to examine associations (pertaining to estimates of effect predicted from genetic variants used as instruments) of adiposity with CRC risk, of adiposity with metabolites, of adiposity-associated metabolites with CRC risk, and finally of adiposity with CRC risk adjusted for representative metabolites. In two-sample MR, SNP-exposure and SNP-outcome associations are obtained from different study sources and combined as a ratio to estimate effects of exposures on outcomes (12, 26). Our study aims and assumptions are shown in **Figure 1**.

**Figure 1.**
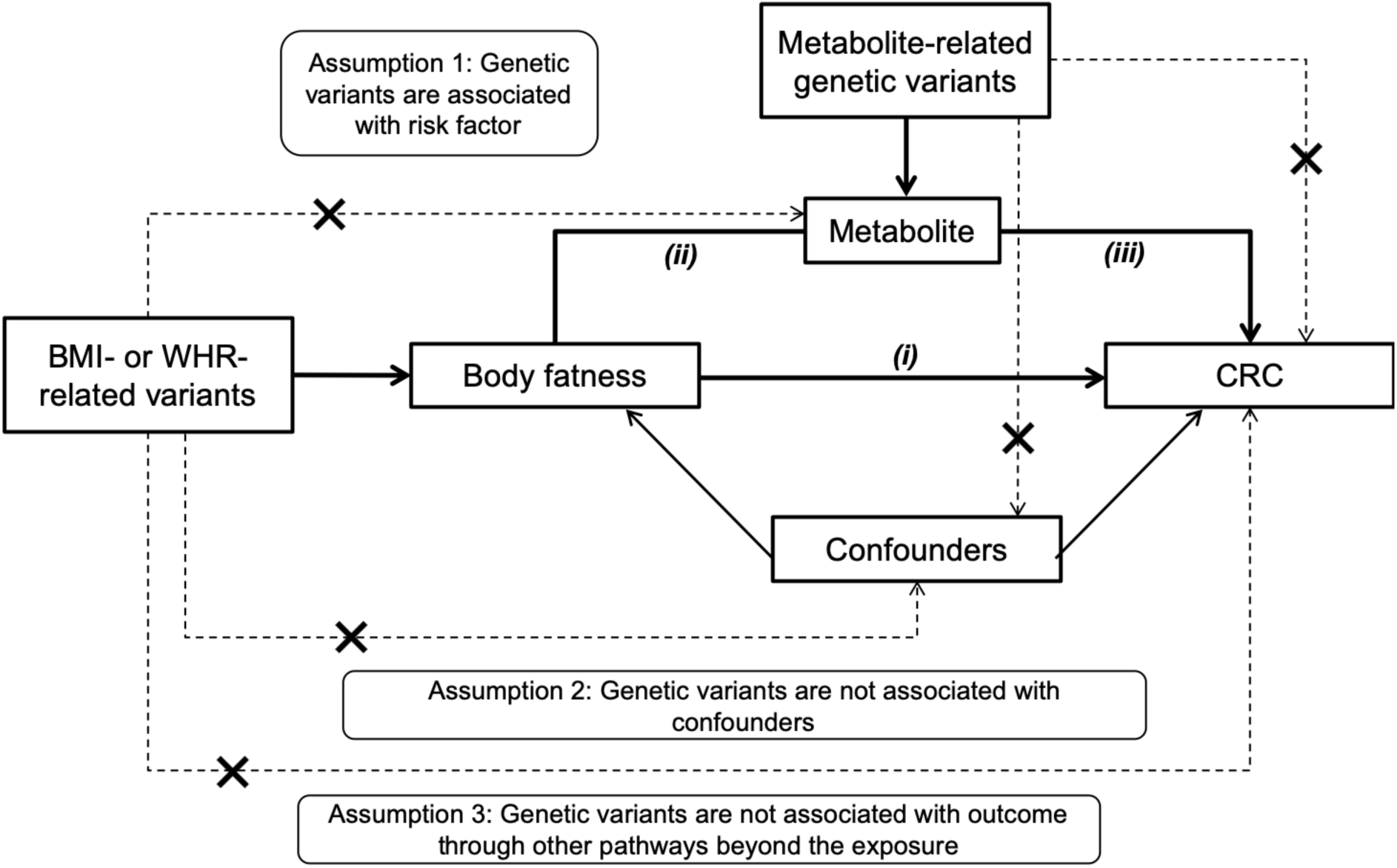
Study aims and assumptions. Study aims are to: 1) estimate the total effect of adiposity on CRC risk using genetic instruments for BMI and WHR ((i), unadjusted for BMI); and 2) estimate the mediated effect of adiposity on CRC risk by metabolites from targeted NMR metabolomics. Aim 2 is addressed using two approaches: 1) two-step MR wherein effects are examined of adiposity on metabolites (ii) and of adiposity-related metabolites on CRC risk (iii); and 2) multivariable MR wherein effects of adiposity on CRC (i) are examined with adjustment for the effect of representative metabolite classes on CRC (iii). Sex specific analyses were performed when sex-specific GWAS estimates for exposure and outcome were both available. When ≥ 2 SNP instruments were available, up to 3 MR models were applied: the inverse variance weighted (IVW) model which assumes that none of the SNPs are pleiotropic (33), the weighted median (WM) model which allows up to half of the included SNPs to be pleiotropic and is less influenced by outliers (33), and the MR-Egger model which provides an estimate of association magnitude allowing all SNPs to be pleiotropic (32). Analyses with metabolites as outcomes were conducted within discovery aims wherein P-value thresholds are applied to prioritize traits with the strongest evidence of association to be taken forward into further stages of analysis (with CRC risk). Analyses with CRC as outcomes were conducted within estimation aims wherein P-values are interpreted as continuous indicators of evidence strength and focus is on effect size and precision (63, 64).

### Adiposity instruments

We identified SNPs that were independently associated (low linkage disequilibrium (LD), R^2^<0.001) with BMI and WHR (unadjusted for BMI) at P<5×10^−8^ from a recent large-scale genome-wide association study (GWAS) meta-analysis of 221,863 to 806,810 male and female adults of European ancestry from the Genetic Investigation of ANthropometric Traits (GIANT) consortium and the UK Biobank (27) (**Supplementary Table 1**). BMI and WHR are expressed in standard deviation (SD) units. For sex-combined analyses of BMI and WHR, 312 and 209 SNPs were used, respectively. For sex-specific analyses of BMI, 185 and 152 SNPs were used for women and men, respectively. For sex-specific analyses of WHR, 153 and 64 SNPs were used for women and men, respectively. F-statistics for adiposity instruments ranged from 75.8 to 124.5 (**Supplementary Table 2**), indicating instrument strength above recommended minimum levels (28).

### Metabolite instruments

We identified SNPs that were independently associated (R^2^<0.001 and P<5×10^−8^) with metabolites from a GWAS of 123 traits from targeted nuclear magnetic resonance (NMR) metabolomics (**Supplementary Table 1**); these included lipoprotein subclass-specific lipids, amino acids, fatty acids, inflammatory glycoproteins, and others (24). Between 13,476 and 24,925 adults (men and women combined) of European ancestry were included. Metabolic traits are expressed in SD units. F-statistics for metabolite instruments ranged from 30.2 to 220.8 (**Supplementary Table 2**), indicating sufficient instrument strength for univariable analyses.

### Colorectal cancer GWAS data

We obtained SNP estimates from the most comprehensive GWAS of CRC to date (25), including 58,221 cases and 67,694 controls from 45 studies within 3 consortia: Genetics and Epidemiology of Colorectal Cancer Consortium (GECCO), Colorectal Cancer Transdisciplinary Study (CORECT) and Colon Cancer Family Registry (CCFR). Cases were diagnosed by a physician and recorded overall and by site (colon, proximal colon, distal colon, rectum). Approximately 92% of participants were white-European (∼8% were East Asian). Case distributions are outlined in **Supplementary Table 3**; other study characteristics are detailed elsewhere (25). Ethics were approved by respective institutional review boards.

### Statistical approach

First, we examined associations of BMI and WHR with overall and site-specific CRC using SNP estimates from sex-combined GWAS. We then examined associations of BMI and WHR with overall CRC based on SNP estimates from sex-specific GWAS (sex-specific GWAS were unavailable for site-specific CRC). As sensitivity analyses, up to three MR models were used which make differing pleiotropy assumptions (detailed in **Figure 1 legend**) (29, 30). When only a single SNP was available, the Wald ratio was used (31). When ≥ 2 SNPs were available, random-effects inverse variance weighted (IVW) (30), MR-Egger (32) and weighted median (WM) (33) models were used. Cochrane’s Q-statistic was used to assess heterogeneity of SNP effects (smaller P-values indicating higher heterogeneity and potential for directional pleiotropy (34)). Scatter plots were used to compare MR models and ‘leave-one-SNP-out’ analyses were used to detect SNP outliers (35).

Second, we examined associations of BMI and WHR with metabolites using results from sex-combined GWAS (sex-specific GWAS were unavailable for metabolites) and the MR models described above. Each metabolite (analysed as an outcome) that was associated with either BMI or WHR based on an IVW model P-value ≤ 0.05 following a false discovery rate (FDR) correction (Benjamini-Hochberg method (36)) was taken forward and examined for association with CRC risk using the IVW model (if ≥ 2 SNPs) or the Wald ratio (if 1 SNP). Multivariable MR (37) was also used to examine associations of BMI and WHR with CRC risk, adjusting for single metabolites that were representative of various metabolite classes based on previous network analyses (38) and that had the highest instrument strength based on the F-statistic (**Supplementary Table 2**). As a positive control, we adjusted BMI for WHR as a covariate (which is expected to attenuate the association of BMI with CRC risk), and likewise we adjusted WHR for BMI as a covariate with the same expectation. A smaller set of SNPs for BMI and WHR based on earlier GWAS (39, 40) was used for these multivariable models to avoid a relative dilution of metabolite instrument strength given that the number of SNPs for BMI and WHR from expanded GWAS far outnumbered those for metabolites.

In each instance, MR estimates are interpreted as the change in outcome per SD-unit change in the exposure. Estimates for metabolite outcomes reflect SD-unit change and estimates for CRC outcomes reflect odds ratios (OR) associated with an SD-unit change in exposure. Statistical analyses were performed using R (Version 3.5.2).

## Results

### Associations of BMI and WHR with CRC risk

In sex-combined analyses (**Figure 2; Supplementary Table 4**), higher BMI (per 4.8 kg/m^2^) was associated with higher risk of overall CRC (IVW-OR=1.16, 95% CI=1.07, 1.26). The WM estimate was similar, but the MR-Egger estimate was reduced (OR=1.02, 95% CI=0.84, 1.25). BMI associations were consistent across CRC sites. Associations were directionally consistent for WHR as for BMI but were marginally stronger – e.g. higher WHR (per 0.09-ratio) was associated with 1.28 (95% CI=1.16, 1.42) times higher odds of CRC in an IVW model (WM and MR-Egger estimates were similar). WHR associations were consistent across CRC sites. SNP heterogeneity was similarly high for BMI and WHR (P-value range across models=9.54×10^−10^ to 1.97×10^−8^).

**Figure 2.**
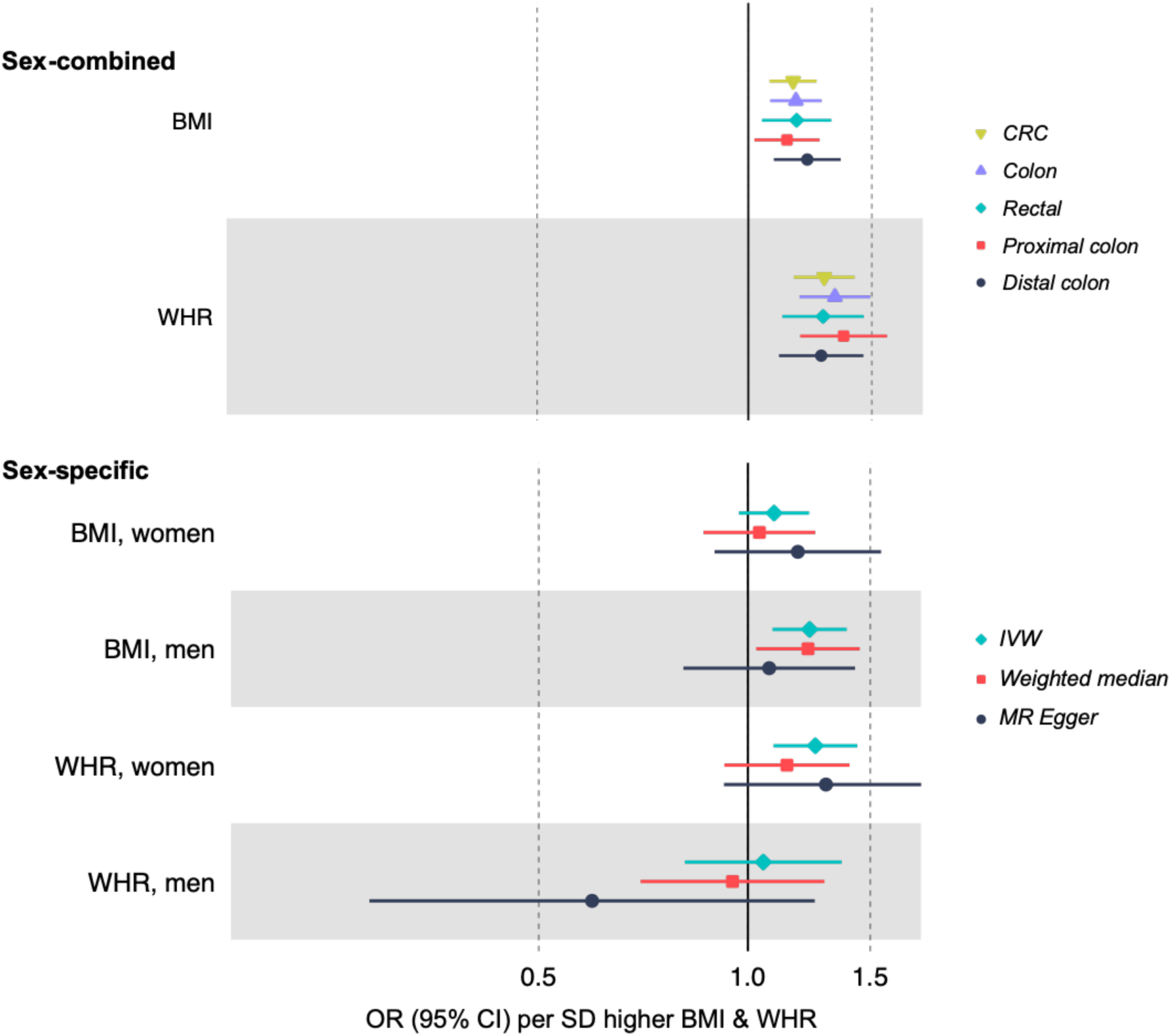
Associations of BMI and WHR with CRC risk based on two-sample MR. Sex-combined estimates are based on GWAS done among women and men together (for both exposure and outcome; IVW models). Sex-specific estimates are based on GWAS done separately among women and men (for both exposure and outcome).

In sex-specific IVW models (**Figure 2; Supplementary Table 4**), higher BMI (per 4.2 kg/m^2^) was associated with 1.23 (95% CI=1.08, 1.38) times higher odds of CRC among men and 1.09 (95% CI=0.97, 1.22) times higher odds of CRC (per 5.2 kg/m^2^) among women. In a WM model, this BMI estimate was robust among men (OR=1.22, 95% CI=1.02, 1.46) but reduced among women (OR=1.04, 95% CI=0.86, 1.26). MR-Egger estimates were similarly imprecise among men and women and SNP heterogeneity was similar for both. In IVW models, higher WHR (per 0.07-ratio) was associated with 1.25 (95% CI=1.08, 1.43) times higher odds of CRC among women; this estimate was 1.05 (95% CI=0.81, 1.36) among men (per 0.07-ratio). This was also supported by WM estimates (OR=1.14, 95% CI=0.91, 1.42 among women and OR=0.95, 95% CI=0.90, 1.29 among men), and MR-Egger estimates. SNP heterogeneity was similarly high among men and women.

Scatter plots comparing different MR models and results of ‘leave-one-SNP-out’ analyses are presented in **Supplemental Figures 1-42**.

### Associations of BMI and WHR with metabolites

In sex-combined analyses, higher BMI (per 4.8 kg/m^2^) or WHR (per 0.09-ratio) was associated with 104 metabolites based on FDR-corrected P-value ≤ 0.05 in IVW models (**Supplementary Figures 43-47; Supplementary Table 5**). Evidence was strong in relation to lipids including total cholesterol and triglycerides in very-low-density lipoproteins (VLDL), low-density lipoproteins (LDL), and high-density lipoproteins (HDL) – e.g. 0.23 SD (95% CI=0.15, 0.31) higher triglycerides in large VLDL from higher BMI. Associations of higher BMI were also strong with lactate, pyruvate, and branched chain amino acids – e.g. 0.19 SD (95% CI=0.13, 0.25) higher isoleucine, and with inflammatory glycoproteins (0.28 SD, 95% CI=0.20, 0.36 higher). Similar patterns were seen for WHR.

### Associations of BMI- or WHR-related metabolites with CRC

Of 104 metabolites associated (as outcomes) with BMI or WHR in sex-combined analyses, 100 had SNPs for use in Wald or IVW models. As shown in **Supplementary Table 1**, 321 unique SNPs were used to instrument 100 metabolites (3 metabolites had 1 SNP, 13 metabolites had < 5 SNPs, and 51 metabolites had < 10 SNPs; SNP counts across metabolites ranged from 1 to 26). Lipid traits showed generally weak associations with CRC which were also in directions inconsistent with mediation of the adiposity-CRC relationship – e.g. lipids in medium HDL were positively associated with CRC, but these had been negatively associated with BMI or WHR (**Figure 3; Supplementary Table 6**). In contrast, there was more consistent evidence of a positive association of lipids in intermediate-density lipoprotein (IDL), VLDL, and LDL with risk of distal colon cancer, and these lipids had been positively associated with higher BMI or WHR. For example, higher total lipids in IDL (per SD) were associated with 1.09 (95% CI=1.02, 1.15) times higher odds of distal colon cancer. Lipids were unassociated with risk of proximal colon cancer. Fatty acids were unassociated with CRC risk except for higher monounsaturated fatty acid levels which were associated with lower risk of rectal cancer (IVW-OR=0.85, 95% CI=0.75, 0.95; **Figure 4**). Lactate and pyruvate were inversely associated with CRC at 0.66 (95% CI=0.42, 1.03) times lower odds and 0.64 (95% CI=0.52, 0.80) times lower odds, respectively. However, these metabolites were positively associated with BMI and so directions were inconsistent with mediation of the adiposity-CRC relationship. Amino acids and glycoprotein acetyls were unassociated with CRC risk.

**Figure 3.**
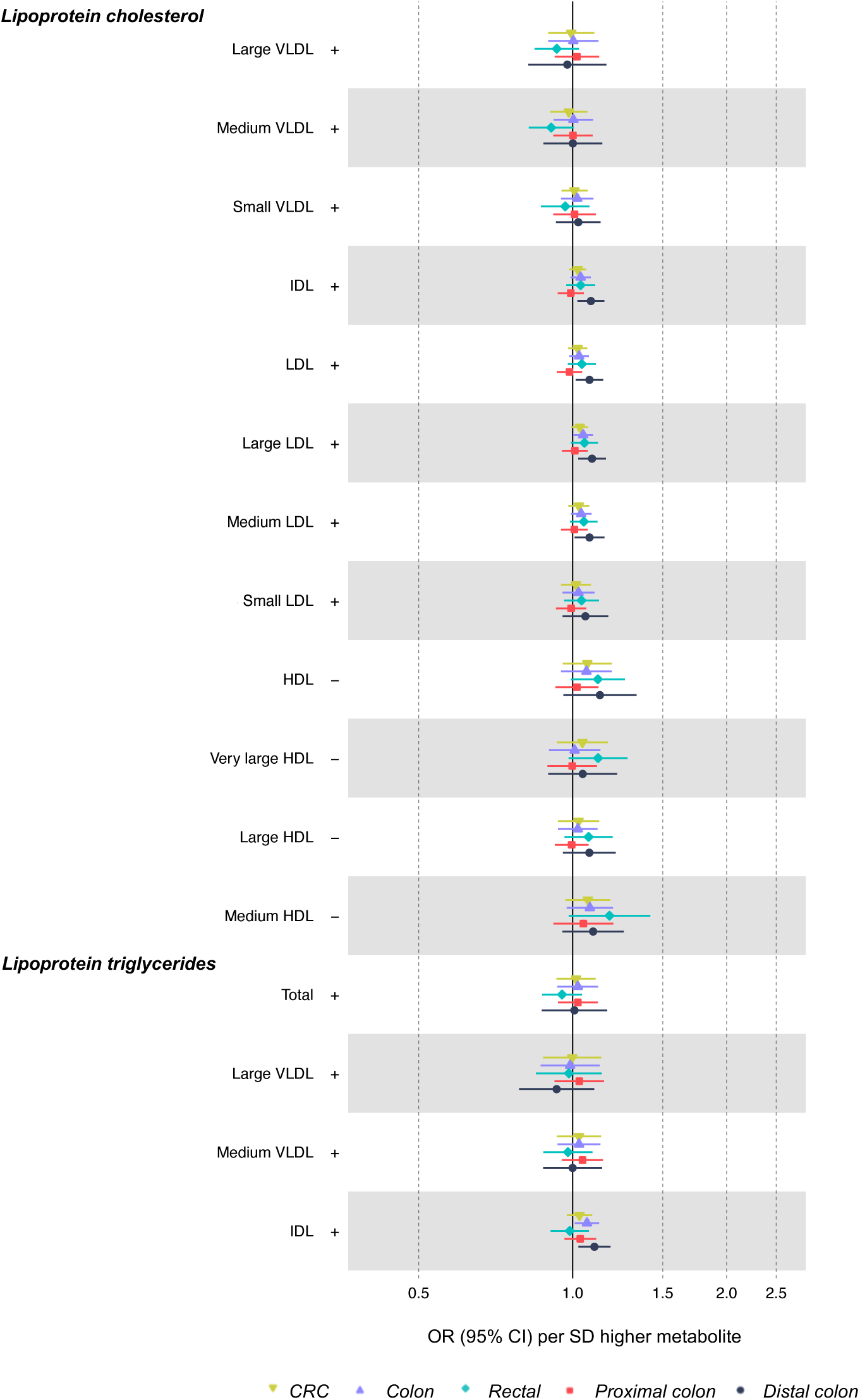
Associations of BMI- or WHR-related lipid metabolites with CRC risk based on two-sample MR (IVW method) Estimates reflect the OR (95% CI) for CRC per SD higher metabolite that is associated (as an outcome) with BMI or WHR. +/- symbols indicate the direction of association of BMI or WHR with that metabolite

**Figure 4.**
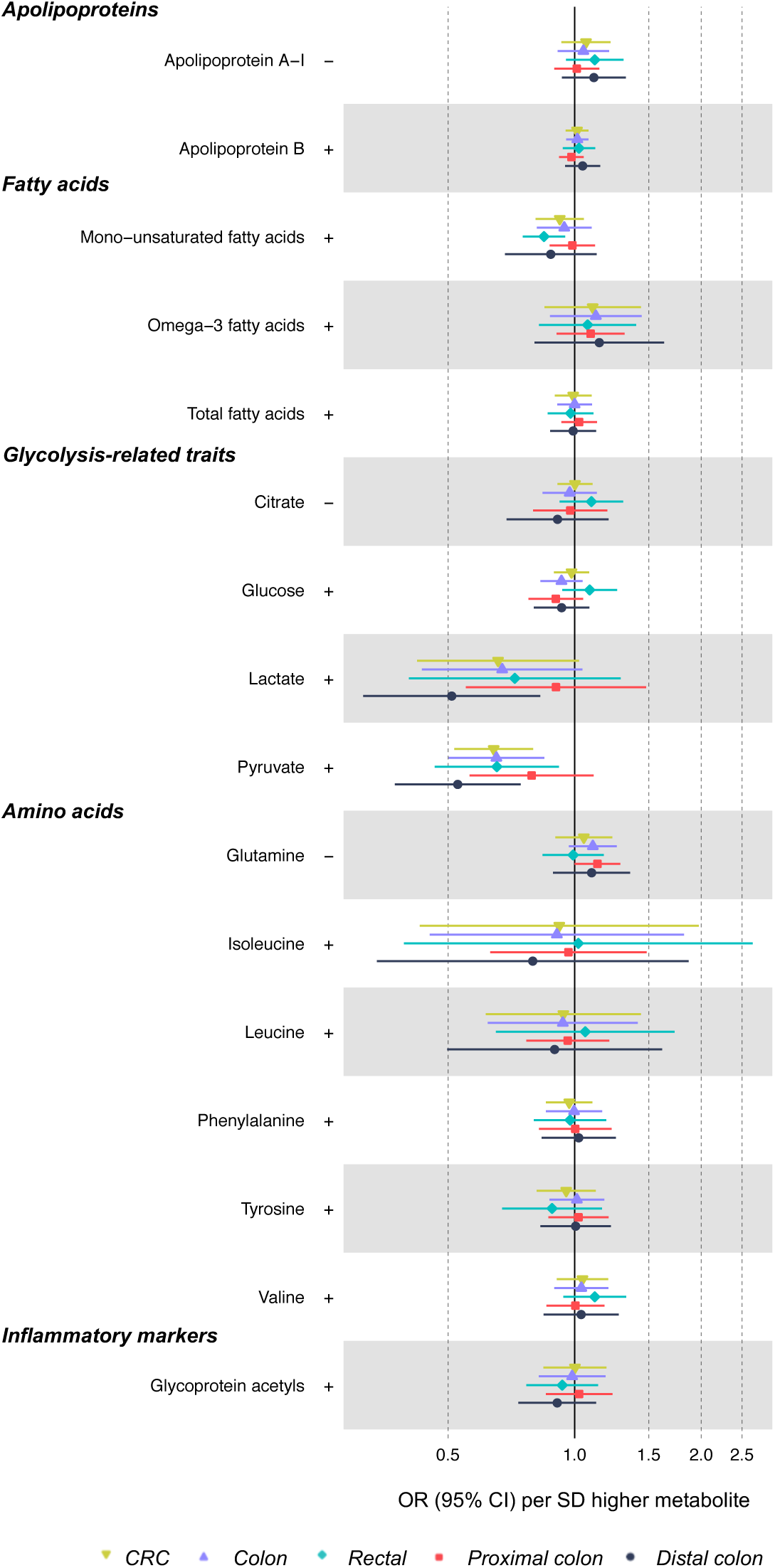
Associations of BMI- or WHR-related non-lipid metabolites with CRC risk based on two-sample MR (IVW method) Estimates reflect the OR (95% CI) for CRC per SD higher metabolite that is associated (as an outcome) with BMI or WHR. +/- symbols indicate the direction of association of BMI or WHR with that metabolite

### Associations of BMI and WHR with CRC risk independent of metabolites

The association of BMI with overall CRC was not attenuated following adjustment for various metabolite classes (**Figure 5; Supplementary Table 7**). The univariable IVW-OR for BMI (per 4.77 kg/m^2^ higher, based on 67 SNPs) in relation to CRC was 1.12 (95% CI=1.00, 1.26), whereas this IVW-OR was 1.14 (95% CI=1.01, 1.29) adjusting for VLDL lipids and 1.11 (95% CI=0.99, 1.26) adjusting for IDL and LDL lipids. Attenuation was greater when adjusting the BMI-CRC association for WHR (positive control), at IVW-OR=0.93 (95% CI=0.78, 1.11). Results for WHR in relation to CRC were directionally consistent as seen for BMI, with a lack of attenuation upon adjustment for metabolite classes.

**Figure 5.**
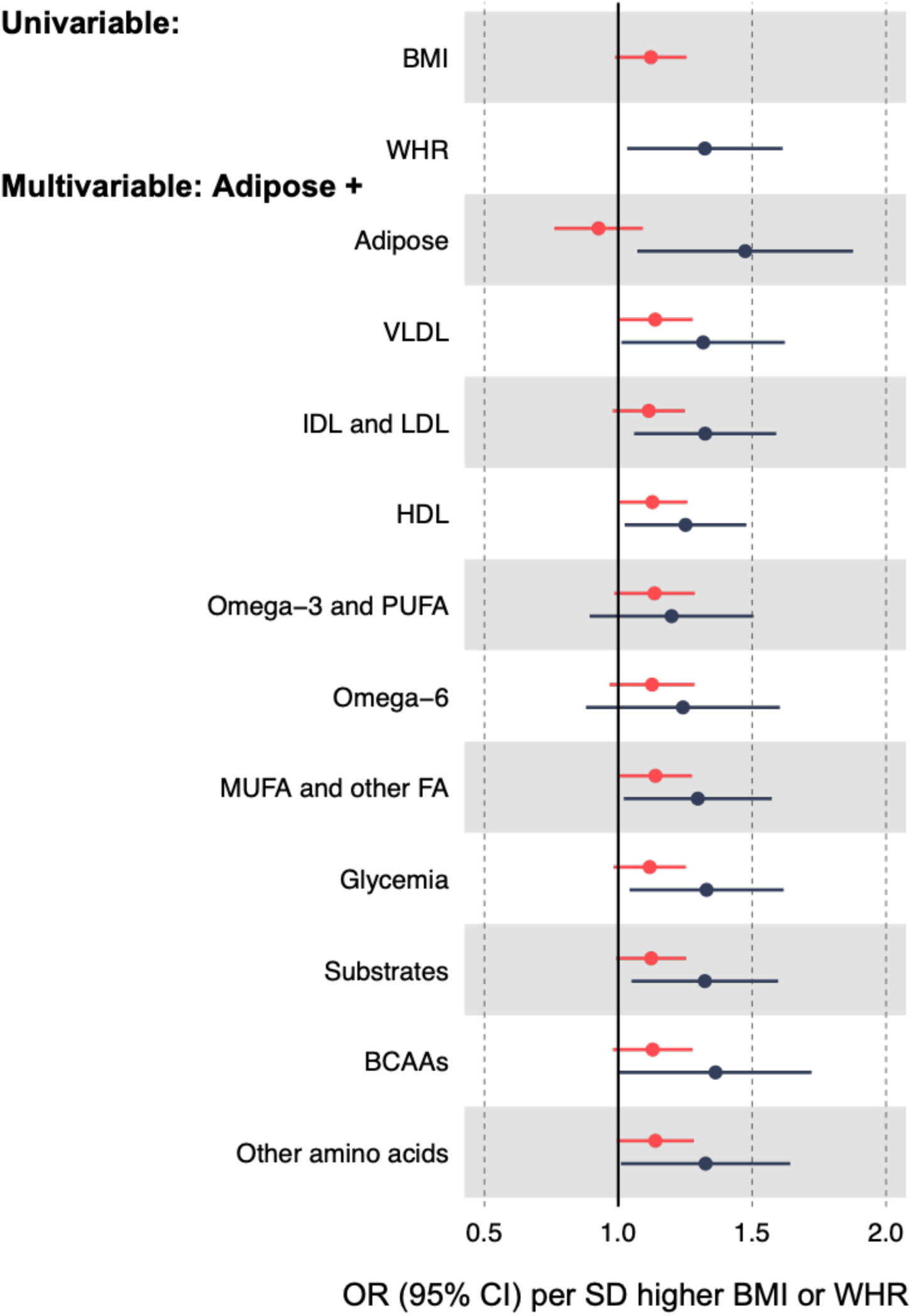
Associations of BMI and WHR with CRC risk independent of various metabolite classes based on multivariable MR. Metabolite classes are based on a single representative metabolite from a previous network analysis (38), as follows: VLDL (triglycerides in small VLDL); IDL and LDL (total cholesterol in medium LDL); HDL (triglycerides in very large HDL); Omega-3 and PUFA (other polyunsaturated fatty acids than 18:2); Omega-6 (18:2, linoleic acid); MUFA and other fatty acids (Omega-9 and saturated fatty acids); glycemia (glucose); substrates (citrate); branched chain amino acids (leucine); other amino acids (glutamine). Adipose adjustments include the alternative adiposity trait (WHR or BMI) as a positive control.

## Discussion

We aimed to better estimate sex-specific effects of adiposity on CRC risk, and to identify potential metabolic mediators of effects of adiposity on CRC, using two-sample MR methods and expanded sample sizes. Our results, based on genetic instruments for adiposity that were sex-specific and were not mutually conditioned, suggest that higher BMI more greatly raises CRC risk among men, whereas higher WHR more greatly raises CRC risk among women. In sex-combined mediation analyses, adiposity was associated with numerous metabolic alterations, but none of these alterations explained associations between adiposity and CRC. More detailed metabolomic measures are likely needed to clarify mechanistic pathways.

Observational (3, 41) and MR studies (6-8) have suggested adverse effects of adiposity on CRC risk, but causal evidence has been lacking regarding sex-specificity. Previous MR studies suggested stronger effects of BMI on CRC risk among women (6-8), which contradicts observational suggestions of stronger effects among men (11). Our new results, based on sex-specific instruments for BMI and WHR and a six-times larger sample size than used previously, suggest that BMI more greatly raises CRC risk among men – a reversal of previous estimates. This new pattern for BMI and CRC (22% higher risk among men per 4.2 kg/m^2^ and 9% higher risk among women per 5.2 kg/m^2^) is highly consistent with observational estimates reviewed by IARC (22% higher risk in men and 9% higher risk in women per 5 kg/m^2^ (6)). Our results also support a reversal of previous estimates for WHR, with risk now appearing higher among women than men. This is unexpected since BMI and abdominal fat measures correlate highly (42, 43); however, given that fat storage is more peripheral in women (17, 18), WHR (unadjusted for BMI) may be a better proxy for extremeness of fat volume among women since fat may be stored more abdominally only when peripheral fat stores are overwhelmed. SNP heterogeneity was high for BMI and WHR with CRC, although this was similar between sexes and directions of effect from sensitivity models were consistent, suggesting balanced SNP heterogeneity.

Given the difficulty of weight loss (10) and the on-going obesity epidemic, it is increasingly important to identify the biological pathways which explain the effect of adiposity on risk of chronic diseases including CRC (9). Adipose tissue is highly metabolically active and secretes pro-inflammatory cytokines such as interleukin (IL)-6 and tumour necrosis factor (TNF)-alpha which may promote tumour initiation (44). Adipose tissue-derived inflammation also promotes insulin resistance in glucose storage tissues that can lead to hyperinsulinemia (45), and insulin and insulin-like growth factors (IGF) such as IGF-1 have pro-mitogenic and anti-apoptotic effects that are cancer promotive (41, 46-50). Our current results suggest effects of BMI or WHR on numerous lipids and pre-glycemic traits; however, few of these traits had any strong association with CRC risk, and the few that did were in a direction that was inconsistent with a mediating role in the adiposity-CRC relationship. Results of a series of multivariable MR models, which adjusted for various metabolites considered representative of broader metabolite classes (37), suggested that associations of BMI and WHR with CRC risk were highly independent of these metabolites. However, this analysis may be limited by weak instrument bias (51) given that F-statistics for metabolite instruments included in each multivariable MR model were relatively low. Nevertheless, results of two complementary approaches to mediation (two-step MR and multivariable MR) provide little evidence that effects of adiposity on CRC risk are mediated by adiposity-related metabolites that are detectable by NMR metabolomics. Future studies could examine metabolites, proteins, hormones, and inflammatory factors that are detectable by other metabolomic and proteomic platforms.

The few traits that did show consistent directions of effect included total lipids in IDL, LDL, and VLDL particles which were raised by BMI and which in turn raised risk of distal colon cancer specifically (not proximal colon or rectal cancer). If robust, this pattern may reflect differential sensitivity of colon regions to lipid exposure owing to divergent functions (the distal colon functions primarily in storage of resultant faecal matter whereas the proximal colon functions primarily in water absorption and faecal solidification (52)), or it may reflect differential detectability through screening (proximal colon tumours tend to be detected in older ages and at more advanced stages (52)). Colorectal anatomical regions may also have distinct molecular features (53); e.g. the distal colon may be more susceptible to p53 mutations and chromosomal instability (54), whereas proximal colon may be more mucinous and susceptible to microsatellite instability and B-Raf proto-oncogene expression (55, 56). Several meta-analyses of long-term follow-ups of randomised controlled trials of LDL cholesterol-lowering statin use suggested no strong evidence of a protective effect of stain use on CRC risk (57-59); CRC sub-sites were largely unexamined. One previous MR study suggested an adverse effect of higher LDL cholesterol, and a protective effect of genetically proxied statin use, on overall CRC risk (20); again, CRC sub-sites were not examined. Observational evidence for LDL cholesterol and CRC risk is less consistent than for total cholesterol or triglycerides (60), but prospective estimates of lipoprotein subclass measures from metabolomic platforms are lacking as these are only recently available at scale.

The limitations of this study include the non-specificity of genetic variants used as instruments for some metabolites which stems from their expectedly correlated nature (e.g. rs1260326, a SNP in *GCKR*, was included in genetic instruments for 54 metabolites). A total of 321 unique SNPs was used to instrument 100 metabolites, but the number of instruments available for a given metabolite was typically small. This limits causal inference for individual traits but should not prevent identification of relevant classes of traits (e.g. lipid, amino acid). It should also be stressed that genetic variants used for metabolites may alter enzyme expression and so serve as instruments for the metabolising enzyme itself, not factors influenced downstream of that enzyme. Since inference in MR applies to the most proximal trait that the genetic variant relates to (14), directing inference to specific glycolytic traits as distinct from their downstream consequences like insulin resistance (61) (a key result of higher fatness and trigger of tumorigenesis (53)) is difficult and requires stronger genetic instruments alongside mechanistic insights from preclinical studies (62). Adiposity was measured indirectly using BMI and WHR because these correlate highly with more objectively measured fat indexes (42, 43) and allow much larger GWAS sample sizes than otherwise possible (comparably strong GWAS were unavailable for waist circumference). Our sex-specific MR investigations were confined to effects of adiposity on overall CRC because sex-specific GWAS were unavailable for site-specific CRC and metabolite outcomes. Sex-stratified GWAS of such outcomes would enable these in future.

Our results based on sex-specific MR instruments and expanded sample sizes suggest that higher BMI more greatly raises CRC risk among men, whereas higher WHR more greatly raises CRC risk among women. In sex-combined mediation analyses, adiposity was associated with numerous metabolic alterations, but none of these alterations explained associations between adiposity and CRC. More detailed metabolomic measures are likely needed to clarify mechanistic pathways.

## Data Availability

The summary-level GWAS data on outcomes used in this study are available following an application to the Genetics and Epidemiology of Colorectal Cancer Consortium (GECCO).

https://www.fredhutch.org/en/research/divisions/public-health-sciences-division/research/cancer-prevention/genetics-epidemiology-colorectal-cancer-consortium-gecco.html

## Consortia funding

Genetics and Epidemiology of Colorectal Cancer Consortium (GECCO): National Cancer Institute, National Institutes of Health, U.S. Department of Health and Human Services (U01 CA164930, U01 CA137088, R01 CA059045, R21 CA191312).

ASTERISK: a Hospital Clinical Research Program (PHRC-BRD09/C) from the University Hospital Center of Nantes (CHU de Nantes) and supported by the Regional Council of Pays de la Loire, the Groupement des Entreprises Françaises dans la Lutte contre le Cancer (GEFLUC), the Association Anne de Bretagne Génétique and the Ligue Régionale Contre le Cancer (LRCC).

The ATBC Study is supported by the Intramural Research Program of the U.S. National Cancer Institute, National Institutes of Health, and by U.S. Public Health Service contract HHSN261201500005C from the National Cancer Institute, Department of Health and Human Services.

CLUE II: This research was funded by the American Institute for Cancer Research and the Maryland Cigarette Restitution Fund at Johns Hopkins, the NCI (U01 CA86308, P30 CA006973 to W.G. Nelson) and the National Institute on Aging (U01 AG18033).

COLO2&3: National Institutes of Health (R01 CA60987).

ColoCare: This work was supported by the National Institutes of Health (grant numbers R01 CA189184 (Li/Ulrich), U01 CA206110 (Ulrich/Li/Siegel/Figueireido/Colditz, 2P30CA015704-40 (Gilliland), R01 CA207371 (Ulrich/Li)), the Matthias Lackas-Foundation, the German Consortium for Translational Cancer Research, and the EU TRANSCAN initiative.

The Colon Cancer Family Registry (CCFR, www.coloncfr.org) was supported in part by funding from the National Cancer Institute (NCI), National Institutes of Health (NIH) (award U01 CA167551) and through U01/U24 cooperative agreements from NCI with the following CCFR centers: Australasian (CA074778 and CA097735), Ontario (OFCCR) (CA074783), Seattle (SFCCR) (CA074794 (and R01 CA076366 to PAN)), USC Consortium (CA074799), Mayo Clinic (CA074800), and Hawaii (CA074806). Support for case ascertainment was provided in part from the Surveillance, Epidemiology, and End Results (SEER) Program and the following U.S. state cancer registries: AZ, CO, MN, NC, NH; and by the Victoria Cancer Registry (Australia) and Ontario Cancer Registry (Canada). Additional funding for the OFCCR/ARCTIC was through award GL201-043 from the Ontario Research Fund (to BWZ), award 112746 from the Canadian Institutes of Health Research (to TJH), through a Cancer Risk Evaluation (CaRE) Program grant from the Canadian Cancer Society (to SG), and through generous support from the Ontario Ministry of Research and Innovation. The SCCFR Illumina HumanCytoSNP array was supported through NCI award R01 CA076366 (to PAN). The CCFR Set-1 (Illumina 1M/1M-Duo) and Set-2 (Illumina Omni1-Quad) scans were supported by NIH awards U01 CA122839 and R01 CA143247 (to GC). The CCFR Set-3 (Affymetrix Axiom CORECT Set array) was supported by NIH award U19 CA148107 and R01 CA81488 (to SBG). The CCFR Set-4 (Illumina OncoArray 600K SNP array) was supported by NIH award U19 CA148107 (to SBG) and by the Center for Inherited Disease Research (CIDR), which is funded by the NIH to the Johns Hopkins University, contract number HHSN268201200008I. Colon Cancer Family Registry (CCFR): The content of this manuscript does not necessarily reflect the views or policies of the NIH or any of the collaborating centers in the CCFR, nor does mention of trade names, commercial products, or organizations imply endorsement by the US Government, any cancer registry, or the CCFR.

COLON: The COLON study is sponsored by Wereld Kanker Onderzoek Fonds, including funds from grant 2014/1179 as part of the World Cancer Research Fund International Regular Grant Programme, by Alpe d’Huzes and the Dutch Cancer Society (UM 2012–5653, UW 2013-5927, UW2015-7946), and by TRANSCAN (JTC2012-MetaboCCC, JTC2013-FOCUS). The Nqplus study is sponsored by a ZonMW investment grant (98-10030); by PREVIEW, the project PREVention of diabetes through lifestyle intervention and population studies in Europe and around the World (PREVIEW) project which received funding from the European Union Seventh Framework Programme (FP7/2007–2013) under grant no. 312057; by funds from TI Food and Nutrition (cardiovascular health theme), a public–private partnership on precompetitive research in food and nutrition; and by FOODBALL, the Food Biomarker Alliance, a project from JPI Healthy Diet for a Healthy Life.

Colorectal Cancer Transdisciplinary (CORECT) Study: The CORECT Study was supported by the National Cancer Institute, National Institutes of Health (NCI/NIH), U.S. Department of Health and Human Services (grant numbers U19 CA148107, R01 CA81488, P30 CA014089, R01 CA197350,; P01 CA196569; R01 CA201407) and National Institutes of Environmental Health Sciences, National Institutes of Health (grant number T32 ES013678).

CORSA: “Österreichische Nationalbank Jubiläumsfondsprojekt” (12511) and Austrian Research Funding Agency (FFG) grant 829675.

CPS-II: The American Cancer Society funds the creation, maintenance, and updating of the Cancer Prevention Study-II (CPS-II) cohort. This study was conducted with Institutional Review Board approval.

CRCGEN: Colorectal Cancer Genetics & Genomics, Spanish study was supported by Instituto de Salud Carlos III, co-funded by FEDER funds –a way to build Europe– (grants PI14-613 and PI09-1286), Agency for Management of University and Research Grants (AGAUR) of the Catalan Government (grant 2017SGR723), and Junta de Castilla y León (grant LE22A10-2). Sample collection of this work was supported by the Xarxa de Bancs de Tumors de Catalunya sponsored by Pla Director d’Oncología de Catalunya (XBTC), Plataforma Biobancos PT13/0010/0013 and ICOBIOBANC, sponsored by the Catalan Institute of Oncology.

Czech Republic CCS: This work was supported by the Grant Agency of the Czech Republic (grants CZ GA CR: GAP304/10/1286 and 1585) and by the Grant Agency of the Ministry of Health of the Czech Republic (grants AZV 15-27580A, AZV 17-30920A and NV18/03/00199).

DACHS: This work was supported by the German Research Council (BR 1704/6-1, BR 1704/6-3, BR 1704/6-4, CH 117/1-1, HO 5117/2-1, HE 5998/2-1, KL 2354/3-1, RO 2270/8-1 and BR 1704/17-1), the Interdisciplinary Research Program of the National Center for Tumor Diseases (NCT), Germany, and the German Federal Ministry of Education and Research (01KH0404, 01ER0814, 01ER0815, 01ER1505A and 01ER1505B).

DALS: National Institutes of Health (R01 CA48998 to M. L. Slattery).

EDRN: This work is funded and supported by the NCI, EDRN Grant (U01 CA 84968-06).

EPIC: The coordination of EPIC is financially supported by the European Commission (DGSANCO) and the International Agency for Research on Cancer. The national cohorts are supported by Danish Cancer Society (Denmark); Ligue Contre le Cancer, Institut Gustave Roussy, Mutuelle Générale de l’Education Nationale, Institut National de la Santé et de la Recherche Médicale (INSERM) (France); German Cancer Aid, German Cancer Research Center (DKFZ), Federal Ministry of Education and Research (BMBF), Deutsche Krebshilfe, Deutsches Krebsforschungszentrum and Federal Ministry of Education and Research (Germany); the Hellenic Health Foundation (Greece); Associazione Italiana per la Ricerca sul Cancro-AIRCItaly and National Research Council (Italy); Dutch Ministry of Public Health, Welfare and Sports (VWS), Netherlands Cancer Registry (NKR), LK Research Funds, Dutch Prevention Funds, Dutch ZON (Zorg Onderzoek Nederland), World Cancer Research Fund (WCRF), Statistics Netherlands (The Netherlands); ERC-2009-AdG 232997 and Nordforsk, Nordic Centre of Excellence programme on Food, Nutrition and Health (Norway); Health Research Fund (FIS), PI13/00061 to Granada, PI13/01162 to EPIC-Murcia, Regional Governments of Andalucía, Asturias, Basque Country, Murcia and Navarra, ISCIII RETIC (RD06/0020) (Spain); Swedish Cancer Society, Swedish Research Council and County Councils of Skåne and Västerbotten (Sweden); Cancer Research UK (14136 to EPIC-Norfolk; C570/A16491 and C8221/A19170 to EPIC-Oxford), Medical Research Council (1000143 to EPIC-Norfolk, MR/M012190/1 to EPICOxford) (United Kingdom).

EPICOLON: This work was supported by grants from Fondo de Investigación Sanitaria/FEDER (PI08/0024, PI08/1276, PS09/02368, P111/00219, PI11/00681, PI14/00173, PI14/00230, PI17/00509, 17/00878, Acción Transversal de Cáncer), Xunta de Galicia (PGIDIT07PXIB9101209PR), Ministerio de Economia y Competitividad (SAF07-64873, SAF 2010-19273, SAF2014-54453R), Fundación Científica de la Asociación Española contra el Cáncer (GCB13131592CAST), Beca Grupo de Trabajo “Oncología” AEG (Asociación Española de Gastroenterología), Fundación Privada Olga Torres, FP7 CHIBCHA Consortium, Agència de Gestió d’Ajuts Universitaris i de Recerca (AGAUR, Generalitat de Catalunya, 2014SGR135, 2014SGR255, 2017SGR21, 2017SGR653), Catalan Tumour Bank Network (Pla Director d’Oncologia, Generalitat de Catalunya), PERIS (SLT002/16/00398, Generalitat de Catalunya), CERCA Programme (Generalitat de Catalunya) and COST Action BM1206 and CA17118. CIBERehd is funded by the Instituto de Salud Carlos III.

ESTHER/VERDI. This work was supported by grants from the Baden-Württemberg Ministry of Science, Research and Arts and the German Cancer Aid.

Harvard cohorts (HPFS, NHS, PHS): HPFS is supported by the National Institutes of Health (P01 CA055075, UM1 CA167552, U01 CA167552, R01 CA137178, R01 CA151993, R35 CA197735, K07 CA190673, and P50 CA127003), NHS by the National Institutes of Health (R01 CA137178, P01 CA087969, UM1 CA186107, R01 CA151993, R35 CA197735, K07CA190673, and P50 CA127003) and PHS by the National Institutes of Health (R01 CA042182).

Hawaii Adenoma Study: NCI grants R01 CA72520.

HCES-CRC: the Hwasun Cancer Epidemiology Study–Colon and Rectum Cancer (HCES-CRC; grants from Chonnam National University Hwasun Hospital, HCRI15011-1).

Kentucky: This work was supported by the following grant support: Clinical Investigator Award from Damon Runyon Cancer Research Foundation (CI-8); NCI R01CA136726.

LCCS: The Leeds Colorectal Cancer Study was funded by the Food Standards Agency and Cancer Research UK Programme Award (C588/A19167).

MCCS cohort recruitment was funded by VicHealth and Cancer Council Victoria. The MCCS was further supported by Australian NHMRC grants 509348, 209057, 251553 and 504711 and by infrastructure provided by Cancer Council Victoria. Cases and their vital status were ascertained through the Victorian Cancer Registry (VCR) and the Australian Institute of Health and Welfare (AIHW), including the National Death Index and the Australian Cancer Database.

MEC: National Institutes of Health (R37 CA54281, P01 CA033619, and R01 CA063464).

MECC: This work was supported by the National Institutes of Health, U.S. Department of Health and Human Services (R01 CA81488 to SBG and GR).

MSKCC: The work at Sloan Kettering in New York was supported by the Robert and Kate Niehaus Center for Inherited Cancer Genomics and the Romeo Milio Foundation. Moffitt: This work was supported by funding from the National Institutes of Health (grant numbers R01 CA189184, P30 CA076292), Florida Department of Health Bankhead-Coley Grant 09BN-13, and the University of South Florida Oehler Foundation. Moffitt contributions were supported in part by the Total Cancer Care Initiative, Collaborative Data Services Core, and Tissue Core at the H. Lee Moffitt Cancer Center & Research Institute, a National Cancer Institute-designated Comprehensive Cancer Center (grant number P30 CA076292).

The Multiethnic Cohort Study is supported by National Cancer Institute grant (CA164973).

NCCCS I & II: We acknowledge funding support for this project from the National Institutes of Health, R01 CA66635 and P30 DK034987.

NFCCR: This work was supported by an Interdisciplinary Health Research Team award from the Canadian Institutes of Health Research (CRT 43821); the National Institutes of Health, U.S. Department of Health and Human Serivces (U01 CA74783); and National Cancer Institute of Canada grants (18223 and 18226). The authors wish to acknowledge the contribution of Alexandre Belisle and the genotyping team of the McGill University and Génome Québec Innovation Centre, Montréal, Canada, for genotyping the Sequenom panel in the NFCCR samples. Funding was provided to Michael O. Woods by the Canadian Cancer Society Research Institute.

NSHDS: Swedish Cancer Society; Cancer Research Foundation in Northern Sweden; Swedish Research Council; J C Kempe Memorial Fund; Faculty of Medicine, Umeå University, Umeå, Sweden; and Cutting-Edge Research Grant from the County Council of Västerbotten, Sweden.

OFCCR: The Ontario Familial Colorectal Cancer Registry was supported in part by the National Cancer Institute (NCI) of the National Institutes of Health (NIH) under award U01 CA167551 and award U01/U24 CA074783 (to SG). Additional funding for the OFCCR and ARCTIC testing and genetic analysis was through and a Canadian Cancer Society CaRE (Cancer Risk Evaluation) program grant and Ontario Research Fund award GL201-043 (to BWZ), through the Canadian Institutes of Health Research award 112746 (to TJH), and through generous support from the Ontario Ministry of Research and Innovation.

OSUMC: OCCPI funding was provided by Pelotonia and HNPCC funding was provided by the NCI (CA16058 and CA67941).

PLCO: Intramural Research Program of the Division of Cancer Epidemiology and Genetics and supported by contracts from the Division of Cancer Prevention, National Cancer Institute, NIH, DHHS. Funding was provided by National Institutes of Health (NIH), Genes, Environment and Health Initiative (GEI) Z01 CP 010200, NIH U01 HG004446, and NIH GEI U01 HG 004438.

SCCFR: The Seattle Colon Cancer Family Registry was supported in part by the National Cancer Institute (NCI) of the National Institutes of Health (NIH) under awards U01 CA167551. Additional support for the SFCCR, Postmenopausal Hormones and Colon Cancer (PMH) study and the SCCFR Illumina HumanCytoSNP array were through NCI/NIH awards U01/U24 CA074794 and R01 CA076366 (to PAN).

SEARCH: The University of Cambridge has received salary support in respect of PDPP from the NHS in the East of England through the Clinical Academic Reserve. Cancer Research UK (C490/A16561); the UK National Institute for Health Research Biomedical Research Centres at the University of Cambridge.

SELECT: Research reported in this publication was supported in part by the National Cancer Institute of the National Institutes of Health under Award Numbers U10 CA37429 (CD Blanke), and UM1 CA182883 (CM Tangen/IM Thompson). The content is solely the responsibility of the authors and does not necessarily represent the official views of the National Institutes of Health.

SMS: This work was supported by the National Cancer Institute (grant P01 CA074184 to J.D.P. and P.A.N., grants R01 CA097325, R03 CA153323, and K05 CA152715 to P.A.N., and the National Center for Advancing Translational Sciences at the National Institutes of Health (grant KL2 TR000421 to A.N.B.-H.)

The Swedish Low-risk Colorectal Cancer Study: The study was supported by grants from the Swedish research council; K2015-55X-22674-01-4, K2008-55X-20157-03-3, K2006-72X-20157-01-2 and the Stockholm County Council (ALF project).

Swedish Mammography Cohort and Cohort of Swedish Men: This work is supported by the Swedish Research Council /Infrastructure grant, the Swedish Cancer Foundation, and the Karolinska Institute’s Distinguished Professor Award to Alicja Wolk.

UK Biobank: This research has been conducted using the UK Biobank Resource under Application Number 8614

VITAL: National Institutes of Health (K05 CA154337).

Women’s Health Initiative: The WHI program is funded by the National Heart, Lung, and Blood Institute, National Institutes of Health, U.S. Department of Health and Human Services through contracts HHSN268201600018C, HHSN268201600001C, HHSN268201600002C, HHSN268201600003C, and HHSN268201600004C.

## Consortia acknowledgements

ASTERISK: We are very grateful to Dr. Bruno Buecher without whom this project would not have existed. We also thank all those who agreed to participate in this study, including the patients and the healthy control persons, as well as all the physicians, technicians and students.

COLON and NQplus: the authors would like to thank the COLON and NQplus investigators at Wageningen University & Research and the involved clinicians in the participating hospitals.

CCFR: The Colon CFR graciously thanks the generous contributions of their 42,505 study participants, dedication of study staff, and the financial support from the U.S. National Cancer Institute, without which this important registry would not exist.

CORSA: We kindly thank all those who contributed to the screening project Burgenland against CRC. Furthermore, we are grateful to Doris Mejri and Monika Hunjadi for laboratory assistance.

CPS-II: The authors thank the CPS-II participants and Study Management Group for their invaluable contributions to this research. The authors would also like to acknowledge the contribution to this study from central cancer registries supported through the Centers for Disease Control and Prevention National Program of Cancer Registries, and cancer registries supported by the National Cancer Institute Surveillance Epidemiology and End Results program.

Czech Republic CCS: We are thankful to all clinicians in major hospitals in the Czech Republic, without whom the study would not be practicable. We are also sincerely grateful to all patients participating in this study.

DACHS: We thank all participants and cooperating clinicians, and Ute Handte-Daub, Utz Benscheid, Muhabbet Celik and Ursula Eilber for excellent technical assistance.

EDRN: We acknowledge all the following contributors to the development of the resource: University of Pittsburgh School of Medicine, Department of Gastroenterology, Hepatology and Nutrition: Lynda Dzubinski; University of Pittsburgh School of Medicine, Department of Pathology: Michelle Bisceglia; and University of Pittsburgh School of Medicine, Department of Biomedical Informatics.

Harvard cohorts (HPFS, NHS, PHS): The study protocol was approved by the institutional review boards of the Brigham and Women’s Hospital and Harvard T.H. Chan School of Public Health, and those of participating registries as required. We would like to thank the participants and staff of the HPFS, NHS and PHS for their valuable contributions as well as the following state cancer registries for their help: AL, AZ, AR, CA, CO, CT, DE, FL, GA, ID, IL, IN, IA, KY, LA, ME, MD, MA, MI, NE, NH, NJ, NY, NC, ND, OH, OK, OR, PA, RI, SC, TN, TX, VA, WA, WY. The authors assume full responsibility for analyses and interpretation of these data.

Kentucky: We would like to acknowledge the staff at the Kentucky Cancer Registry.

LCCS: We acknowledge the contributions of Jennifer Barrett, Robin Waxman, Gillian Smith and Emma Northwood in conducting this study.

NCCCS I & II: We would like to thank the study participants, and the NC Colorectal Cancer Study staff.

PLCO: The authors thank the PLCO Cancer Screening Trial screening center investigators and the staff from Information Management Services Inc and Westat Inc. Most importantly, we thank the study participants for their contributions that made this study possible.

The SCCFR and the PMH study graciously thanks the generous contributions of their study participants, dedication of study staff, and the financial support from the U.S. National Cancer Institute, without which this important research was not possible. The content of this manuscript does not necessarily reflect the views or policies of the NIH or any of the collaborating centers in the CCFR, nor does mention of trade names, commercial products, or organizations imply endorsement by the US Government, any cancer registry, or the CCFR.

SEARCH: We thank the SEARCH team.

SELECT: We thank the research and clinical staff at the sites that participated on SELECT study, without whom the trial would not have been successful. We are also grateful to the 35,533 dedicated men who participated in SELECT.

Women’s Health Initiative: The authors thank the WHI investigators and staff for their dedication, and the study participants for making the program possible. A full listing of WHI investigators can be found at: http://www.whi.org/researchers/Documents%20%20Write%20a%20Paper/WHI%20Investigator%20Short%20List.pdf

## References

1. Sung H, Siegel RL, Rosenberg PS, Jemal A. Emerging cancer trends among young adults in the USA: analysis of a population-based cancer registry. Lancet Public Health. 2019.

2. Mauri G, Sartore-Bianchi A, Russo AG, Marsoni S, Bardelli A, Siena S. Early-onset colorectal cancer in young individuals. Mol Oncol. 2019;13(2):109–31.

3. World Cancer Research Fund/American Institute for Cancer Research. Continuous Update Project Expert Report. Diet, nutrition, physical activity and colorectal cancer. 2018.

4. Lauby-Secretan B, Scoccianti C, Loomis D, Grosse Y, Bianchini F, Straif K. Body Fatness and Cancer—Viewpoint of the IARC Working Group. N Engl J Med. 2016;375(8):794–8.

5. Davey Smith G, Ebrahim S. ‘Mendelian randomization’: can genetic epidemiology contribute to understanding environmental determinants of disease? Int J Epidemiol. 2003;32(1):.

6. Thrift AP, Gong J, Peters U, Chang-Claude J, Rudolph A, Slattery ML, et al. Mendelian randomization study of body mass index and colorectal cancer risk. Cancer Epidemiol Biomarkers Prev. 2015;24(7):1024–31.

7. Jarvis D, Mitchell JS, Law PJ, Palin K, Tuupanen S, Gylfe A, et al. Mendelian randomisation analysis strongly implicates adiposity with risk of developing colorectal cancer. Br J Cancer. 2016;115(2):266–72.

8. Gao C, Patel CJ, Michailidou K, Peters U, Gong J, Schildkraut J, et al. Mendelian randomization study of adiposity-related traits and risk of breast, ovarian, prostate, lung and colorectal cancer. Int J Epidemiol. 2016;45(3):896–908.

9. Gunter MJ, Riboli E. Obesity and gastrointestinal cancers—where do we go from here? Nature Rev Gastroenterol Hepatol. 2018;15(11):651.

10. Dombrowski SU, Knittle K, Avenell A, Araujo-Soares V, Sniehotta FF. Long term maintenance of weight loss with non-surgical interventions in obese adults: systematic review and meta-analyses of randomised controlled trials. BMJ. 2014;348:g2646.

11. World Cancer Research Fund/American Institute for Cancer Research. Diet, nutrition, physical activity and colorectal cancer: Continuous Update Project. 2017.

12. Lawlor DA. Commentary: Two-sample Mendelian randomization: opportunities and challenges. Int J Epidemiol. 2016;45(3):908.

13. Aschard H, Vilhjálmsson BJ, Joshi AD, Price AL, Kraft P. Adjusting for heritable covariates can bias effect estimates in genome-wide association studies. Am J Hum Genet. 2015;96(2):329–39.

14. Holmes MV, Ala-Korpela M, Davey Smith G. Mendelian randomization in cardiometabolic disease: challenges in evaluating causality. Nat Rev Cardiol. 2017;14:577–90.

15. Hartwig FP, Tilling K, Davey-Smith G, Lawlor DA, Borges M-CJB. Bias in two-sample Mendelian randomization by using covariable-adjusted summary associations. 2019:816363.

16. Holmes MV, Davey Smith G. Problems in interpreting and using GWAS of conditional phenotypes illustrated by’alcohol GWAS’. Mol Psych. 2019;24(2):167.

17. Rosen ED, Spiegelman BM. What we talk about when we talk about fat. Cell. 2014;156(1):20–44.

18. Kahn SE, Hull RL, Utzschneider KM. Mechanisms linking obesity to insulin resistance and type 2 diabetes. Nature. 2006;444(7121):840–6.

19. Würtz P, Wang Q, Kangas AJ, Richmond RC, Skarp J, Tiainen M, et al. Metabolic signatures of adiposity in young adults: Mendelian randomization analysis and effects of weight change. PLoS Med. 2014;11(12):e1001765.

20. Rodriguez-Broadbent H, Law PJ, Sud A, Palin K, Tuupanen S, Gylfe A, et al. Mendelian randomisation implicates hyperlipidaemia as a risk factor for colorectal cancer. Int J Cancer. 2017;140(12):2701–8.

21. Song M, Lu Y, Gunter M, Murphy N, Banbury BL, Ma W, et al. Type 2 diabetes and glycemic traits in relation to colorectal cancer risk: A Mendelian randomization study. AACR; 2018.

22. May-Wilson S, Sud A, Law PJ, Palin K, Tuupanen S, Gylfe A, et al. Pro-inflammatory fatty acid profile and colorectal cancer risk: A Mendelian randomisation analysis. Eur J Cancer. 2017;84:228–38.

23. Würtz P, Kangas AJ, Soininen P, Lawlor DA, Davey Smith G, Ala-Korpela M. Quantitative Serum NMR Metabolomics in Large-Scale Epidemiology: A Primer on-Omic Technology. Am J Epidemiol. 2017:kwx016.

24. Kettunen J, Demirkan A, Würtz P, Draisma HH, Haller T, Rawal R, et al. Genome-wide study for circulating metabolites identifies 62 loci and reveals novel systemic effects of LPA. Nat Commun. 2016;7:11122.

25. Huyghe JR, Bien SA, Harrison TA, Kang HM, Chen S, Schmit SL, et al. Discovery of common and rare genetic risk variants for colorectal cancer. Nature Genet. 2019;51(1):76.

26. Davey Smith G, Hemani G. Mendelian randomization: genetic anchors for causal inference in epidemiological studies. Hum Mol Gen. 2014;23(R1):R89–R98.

27. Pulit SL, Stoneman C, Morris AP, Wood AR, Glastonbury CA, Tyrrell J, et al. Meta-analysis of genome-wide association studies for body fat distribution in 694,649 individuals of European ancestry. bioRxiv. 2018:304030.

28. Haycock PC, Burgess S, Wade KH, Bowden J, Relton C, Davey Smith G. Best (but oft-forgotten) practices: the design, analysis, and interpretation of Mendelian randomization studies. Am J Clin Nutr. 2016;103(4):965–78.

29. Burgess S, Bowden J, Fall T, Ingelsson E, Thompson SG. Sensitivity analyses for robust causal inference from Mendelian randomization analyses with multiple genetic variants. Epidemiology. 2017;28(1):30.

30. Hemani G, Zheng J, Elsworth B, Wade KH, Haberland V, Baird D, et al. The MR-Base platform supports systematic causal inference across the human phenome. eLife. 2018;7:e34408.

31. Wald A. The fitting of straight lines if both variables are subject to error. Ann Mathematical Statistics. 1940;11(3):284–300.

32. Bowden J, Davey Smith G, Burgess S. Mendelian randomization with invalid instruments: effect estimation and bias detection through Egger regression. Int J Epidemiol. 2015;44(2):512–25.

33. Bowden J, Davey Smith G, Haycock PC, Burgess S. Consistent estimation in Mendelian randomization with some invalid instruments using a weighted median estimator. Genet Epidemiol. 2016;40(4):304–14.

34. Bowden J, Hemani G, Davey Smith GJAjoe. Invited Commentary: Detecting Individual and Global Horizontal Pleiotropy in Mendelian Randomization—A Job for the Humble Heterogeneity Statistic? 2018;187(12):2681–5.

35. Zheng J, Baird D, Borges M-C, Bowden J, Hemani G, Haycock P, et al. Recent developments in Mendelian randomization studies. Curr Epidemiol Rep. 2017;4(4):330–45.

36. Benjamini Y, Hochberg Y. Controlling the false discovery rate: a practical and powerful approach to multiple testing. J Royal Statistic Soc: Series B (Methodological). 1995;57(1):289–300.

37. Sanderson E, Davey Smith G, Windmeijer F, Bowden J. An examination of multivariable Mendelian randomization in the single-sample and two-sample summary data settings. Int J Epidemiol. 2018;dyy262:1–15.

38. Kujala UM, Mäkinen V-P, Heinonen I, Soininen P, Kangas AJ, Leskinen TH, et al. Long-term leisure-time physical activity and serum metabolome. Circulation. 2012:CIRCULATIONAHA. 112.105551.

39. Locke AE, Kahali B, Berndt SI, Justice AE, Pers TH, Day FR, et al. Genetic studies of body mass index yield new insights for obesity biology. Nature. 2015;518(7538):197–206.

40. Shungin D, Winkler TW, Croteau-Chonka DC, Ferreira T, Locke AE, Mägi R, et al. New genetic loci link adipose and insulin biology to body fat distribution. Nature. 2015;518(7538):187–96.

41. Murphy N, Jenab M, Gunter MJ. Adiposity and gastrointestinal cancers: epidemiology, mechanisms and future directions. Nature Reviews Gastroenterology & Hepatology. 2018;15:659–70.

42. Bell JA, Carslake D, O’Keeffe LM, Frysz M, Howe LD, Hamer M, et al. Associations of Body Mass and Fat Indexes With Cardiometabolic Traits. J Am Coll Cardiol. 2018;72(24):3142–54.

43. Flegal KM, Shepherd JA, Looker AC, Graubard BI, Borrud LG, Ogden CL, et al. Comparisons of percentage body fat, body mass index, waist circumference, and waist-stature ratio in adults. Am J Clin Nutr. 2009;89(2):500–8.

44. Wei H-J, Zeng R, Lu J-H, Lai W-FT, Chen W-H, Liu H-Y, et al. Adipose-derived stem cells promote tumor initiation and accelerate tumor growth by interleukin-6 production. Oncotarget. 2015;6:7713–26.

45. Hotamisligil G. Inflammation and metabolic disorders. Nature. 2006;444(7121):860–7.

46. Rinaldi S, Cleveland R, Norat T, Biessy C, Rohrmann S, Linseisen J, et al. Serum levels of IGF-I, IGFBP-3 and colorectal cancer risk: results from the EPIC cohort, plus a meta-analysis of prospective studies. International Journal of Cancer. 2010;126:NA-NA.

47. Tran TT, Naigamwalla D, Oprescu AI, Lam L, McKeown-Eyssen G, Bruce WR, et al. Hyperinsulinemia, But Not Other Factors Associated with Insulin Resistance, Acutely Enhances Colorectal Epithelial Proliferation *in Vivo*. Endocrinology. 2006;147:1830–7.

48. Kiunga GA, Raju J, Sabljic N, Bajaj G, Good CK, Bird RP. Elevated insulin receptor protein expression in experimentally induced colonic tumors. Cancer Letters. 2004;211:145–53.

49. Kaaks R, Toniolo P, Akhmedkhanov A, Lukanova A, Biessy C, Dechaud H, et al. Serum C-peptide, insulin-like growth factor (IGF)-I, IGF-binding proteins, and colorectal cancer risk in women. Journal of the National Cancer Institute. 2000;92:1592–600.

50. Murphy N, Carreras-Torres R, Song M, Chan AT, Martin RM, Papadimitriou N, et al. Circulating Levels of Insulin-like Growth Factor 1 and Insulin-like Growth Factor Binding Protein 3 Associate With Risk of Colorectal Cancer Based on Serologic and Mendelian Randomization Analyses. Gastroenterology. 2019.

51. Burgess S, Thompson SG, epidemiology CCGCJIjo. Avoiding bias from weak instruments in Mendelian randomization studies. 2011;40(3):755–64.

52. Gonzalez EC, Roetzheim RG, Ferrante JM, Campbell R. Predictors of proximal vs. distal colorectal cancers. Diseases of the colon and rectum. 2001;44:251–8.

53. Jacobs ET, Thompson PA, Mart??nez MaE. Diet, Gender, and Colorectal Neoplasia. Journal of Clinical Gastroenterology. 2007;41:731–46.

54. Okubo R, Masuda H, Nemoto N. p53 mutation found to be a significant prognostic indicator in distal colorectal cancer. Oncology reports.8:509–14.

55. Pekow J, Meckel K, Dougherty U, Butun F, Mustafi R, Lim J, et al. Tumor suppressors miR-143 and miR-145 and predicted target proteins API5, ERK5, K-RAS, and IRS-1 are differentially expressed in proximal and distal colon. American Journal of Physiology-Gastrointestinal and Liver Physiology. 2015;308:G179–G87.

56. Missiaglia E, Jacobs B, D’Ario G, Di Narzo AF, Soneson C, Budinska E, et al. Distal and proximal colon cancers differ in terms of molecular, pathological, and clinical features. Annals of Oncology. 2014;25:1995–2001.

57. Dale KM, Coleman CI, Henyan NN, Kluger J, White CM. Statins and cancer risk: a meta-analysis. JAMA. 2006;295(1):74–80.

58. Liu Y, Tang W, Wang J, Xie L, Li T, He Y, et al. Association between statin use and colorectal cancer risk: a meta-analysis of 42 studies. 2014;25(2):237–49.

59. Lytras T, Nikolopoulos G, Bonovas SJWJoGW. Statins and the risk of colorectal cancer: an updated systematic review and meta-analysis of 40 studies. 2014;20(7):1858.

60. Yao X, Tian Z. Dyslipidemia and colorectal cancer risk: a meta-analysis of prospective studies. Cancer Causes & Control. 2015;26(2):257–68.

61. Lee S, Zhang C, Kilicarslan M, Piening BD, Bjornson E, Hallström BM, et al. Integrated network analysis reveals an association between plasma mannose levels and insulin resistance. Cell metabolism. 2016;24(1):172–84.

62. Lawlor DA, Tilling K, Davey Smith G. Triangulation in aetiological epidemiology. International journal of epidemiology. 2016;45(6):1866–86.

63. Sterne JA, Davey Smith G. Sifting the evidence—what’s wrong with significance tests? BMJ. 2001;322(7280):.

64. Wasserstein RL, Lazar NA. The ASA’s statement on p-values: context, process, and purpose. Am Statistician. 2016;70(2):129–33.

